# Heat Exposure, Occupational Injury Risk, and Economic Costs in New York State

**DOI:** 10.64898/2026.04.20.26351297

**Authors:** Zoey Laskaris, Sherry Baron, Steven B. Markowitz

**Affiliations:** Queens College, City University of New York

## Abstract

**Objectives:** Rising temperatures are a major climate-related hazard for U.S. workers, increasing heat-related illness and a broad range of occupational injuries through indirect pathways often overlooked in economic evaluations. We examined the association between temperature and occupational injury and illness and quantified heat-attributable injuries (including illnesses) and costs in New York State.

**Methods:** We conducted a time-stratified case-crossover study of 591,257 workers’ compensation (WC) claims during the warm season (2016–2024). Daily maximum temperature was linked to injury date and county and modeled using natural cubic splines, with effect modification by industry and worker characteristics.

**Results:** Injury risk increased with temperature, becoming statistically significant at approximately 78°F. Relative to 65°F, injury odds increased to 1.06 (95% CI: 1.01–1.10) at 80°F, 1.12 (1.07–1.18) at 90°F, and 1.17 (1.11–1.23) at 95°F. Overall, 5.0% of claims (2,322 annually) were attributable to heat. At temperatures ≥80°F, an estimated 1,729 excess injuries occurred annually, generating approximately $46 million in WC costs. An estimated $3.2 million to $36.1 million in medical expenditures were associated with incomplete claims, likely borne outside the WC system.

**Conclusions:** These findings demonstrate substantial economic costs not fully captured within WC and support workplace heat protections as a cost-containment strategy that can reduce health care spending and strengthen workforce resilience.

## Introduction

Rising temperatures are among the most pressing climate-related hazards threatening labor in the United States (U.S.), affecting an estimated 37 million workers each year.^1-4^ They increase the risk of both heat-related illness and a broader range of occupational injuries not explicitly coded as heat-related through mechanisms including fatigue, impaired cognition, and loss of coordination.^5, 6^

This distinction between heat-coded cases and heat-attributable risk has important implications for measuring the full social and economic impact of heat. Surveillance systems such as the Census of Fatal Occupational Injuries (CFOI) and the Survey of Occupational Injuries and Illnesses (SOII) capture only cases explicitly coded as heat-related and are known to underestimate occupational injury burden.^7-10^For example, SOII recorded 6,330 injuries requiring days away from work in 2023–2024 with environmental heat as the source, likely representing only a fraction of true heat-attributable harm.^11^

In contrast, quasi-experimental studies leveraging natural variation in temperature capture both directly coded heat illness and indirectly heat-related injuries. These studies consistently show increases in traumatic occupational injuries on hotter days, translating to tens of thousands of heat-attributable injuries annually.^12-17^

However, the associated economic burden, particularly for indirectly heat-related injuries, remains poorly characterized.^18-20^ Workers’ compensation (WC) data provide a key but underused source for estimating these costs.

Understanding the harm and costs associated with workplace heat is important, as heat-related injuries and illnesses are largely preventable, yet most U.S. workers remain unprotected.^5^ Only seven states have enforceable heat standards, and a proposed federal standard - estimated to reduce heat-related injuries by 65% and fatalities by 95% - remains uncertain.^1, 21^

New York State (NY) provides an important and policy-relevant setting: it has a large, diverse workforce with substantial heat exposure but no enforceable standard, despite ongoing legislative efforts (e.g., the Temperature Extreme Mitigation Program (TEMP) Act).^22-25^ Existing NY-based evidence is limited and inconsistent, with surveillance data suggesting low rates of heat-related illness, while WC-based evidence from regional or preliminary analyses suggests increased injury risk on hotter days, underscoring the need for methods that capture indirect heat effects.^3, 4, 16, 26, 27^

We address two key gaps: (1) limited evidence on heat and occupational injury and illness risk in a large, Northern state, and (2) dearth of economic burden estimates that include indirect injuries and illnesses. Using WC claims from 2016–2024, we apply a time-stratified case-crossover design to estimate temperature– injury associations, examine differences across worker and injury characteristics, and quantify heat-attributable injuries and illnesses (hereafter, “injuries”) and costs. Because many reported claims do not progress through the WC system and lack recorded expenditures, we also estimate heat-attributable burden among these incomplete claims. These reported injuries may generate costs outside WC, so we present associated costs as a range reflecting potential expenditures borne by workers, private insurers, or public programs.

## Methods

### Data Source and Study Population

We conducted a time-stratified case-crossover study using publicly available workers’ compensation (WC) claims from the New York State Workers’ Compensation Board (NYS WCB), which include both occupational injuries and illnesses, during the warm season (April 1–October 31), 2016–2024.^28^

### Analytic Samples

NYS WCB data comprise three nested claim subsets reflecting successive stages of case processing: assembled (all reported claims for which the Board opened a case file), complete (claims with formal notice of injury from the insurer and qualifying medical documentation), and established (subset of complete claims with a Board issued determination on work-relatedness and employer liability) (Figure 1).

**Figure 1:**
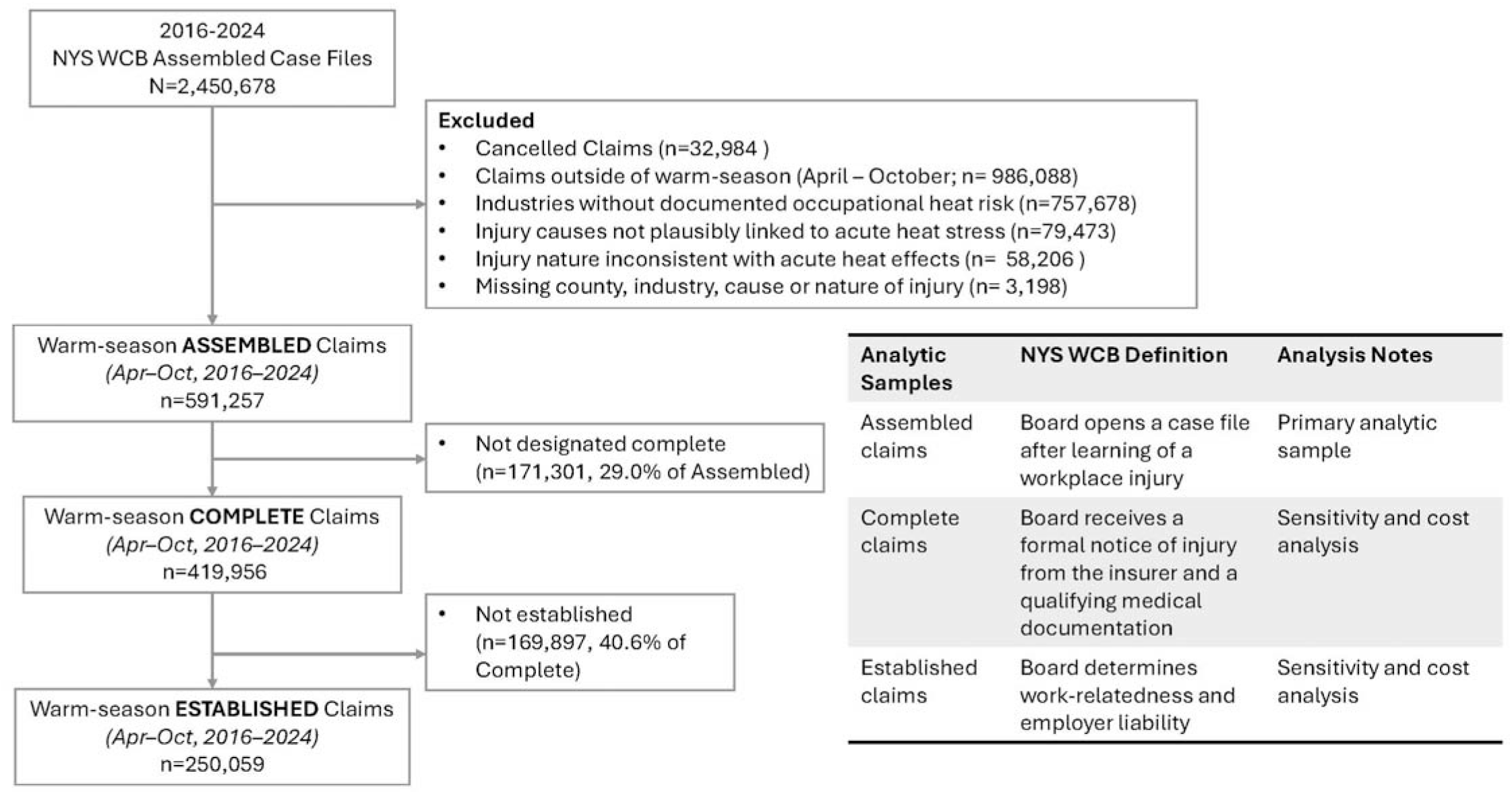
Structure of New York State Workers’ Compensation Claims and Analytic Samples. **Notes:** The NYS Workers’ Compensation Board maintains three nested administrative claim designations reflecting successive stages of case processing: assembled, complete, and established. Warm-season assembled claims (April– October) were used for primary analyses to capture overall injury incidence; complete and established claims were used for sensitivity and cost analyses because they include payment data. Costs for claims not designated as complete were estimated separately. Selection of assembled claims is supported by comparisons with Bureau of Labor Statistics data (see Supplemental Methods and Supplemental Table 1).

Primary analyses used assembled claims to capture the broadest set of reported injuries, which most closely approximated underlying occupational injury incidence (see Supplemental Methods and Supplemental Table 1 for comparisons with BLS injury surveillance data).

Assembled claims that did not progress to complete status (hereafter, incomplete claims) were retained. Although reasons for non-progression are not captured, prior research suggests administrative and legal barriers may affect claim progression; excluding these reported injuries could systematically underestimate injury burden.^29-32^

Complete and established claims were used in sensitivity and cost analyses, because they contain information on insurer-reported costs.

Additional inclusionary and exclusionary criteria are summarized in Figure 1. Analyses were restricted to acute injuries occurring in industries with documented heat exposure risk; cancelled claims and those missing key variables were excluded.

### Injury Cases and Claimant Information

Claimant characteristics included age at the time of injury, gender, length of employment, and industry sector. Injury characteristics were classified using cause and nature of injury codes and restricted to acute injury types plausibly related to short-term heat exposure and acute traumatic or systemic outcomes (see specific causes and nature listed in Table 1 and Supplemental Table 3).

**Table 1:**
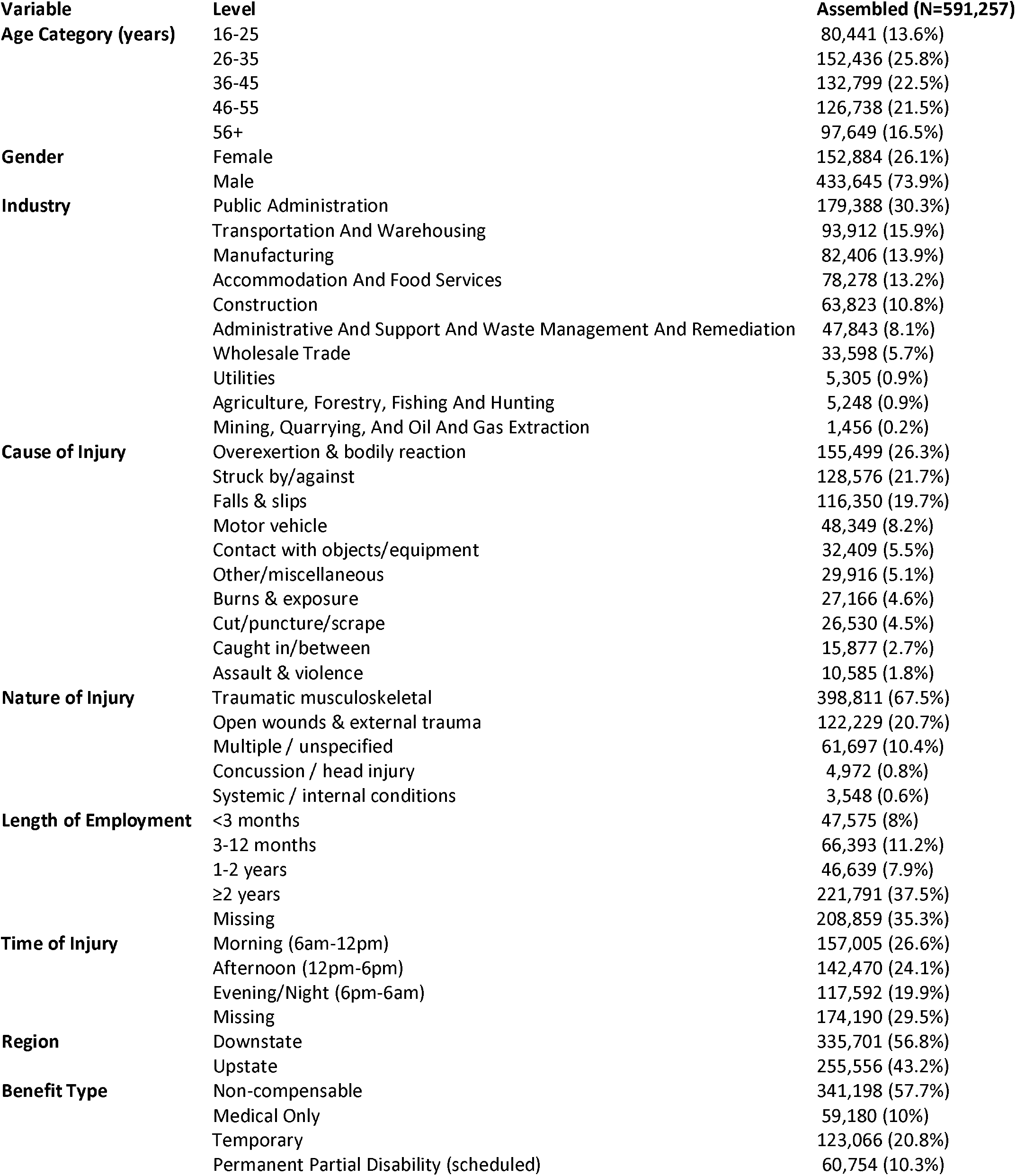
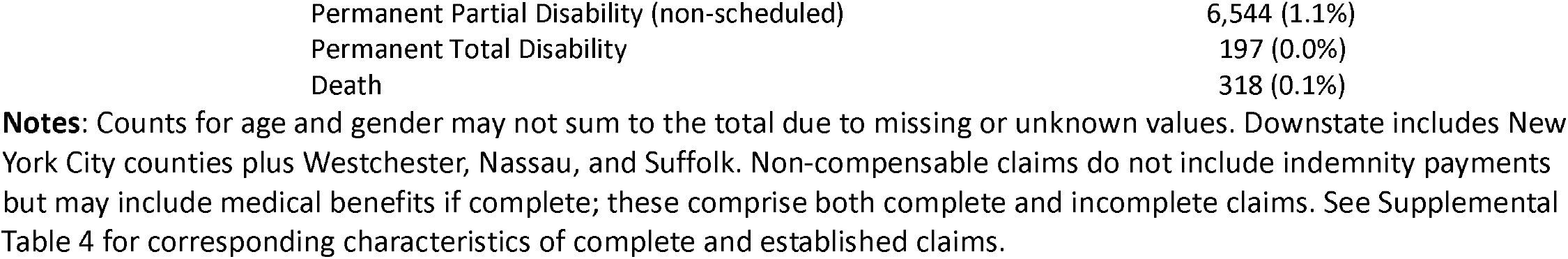
Key descriptive statistics of New York assembled workers’ compensation.

| Variable | Level | Assembled (N=591,257) |
| --- | --- | --- |
| Age Category (years) | 16-25 | 80,441 (13.6%) |
|  | 26-35 | 152,436 (25.8%) |
|  | 36-45 | 132,799 (22.5%) |
|  | 46-55 | 126,738 (21.5%) |
|  | 56+ | 97,649 (16.5%) |
| Gender | Female | 152,884 (26.1%) |
|  | Male | 433,645 (73.9%) |
| Industry | Public Administration | 179,388 (30.3%) |
|  | Transportation And Warehousing | 93,912 (15.9%) |
|  | Manufacturing | 82,406 (13.9%) |
|  | Accommodation And Food Services | 78,278 (13.2%) |
|  | Construction | 63,823 (10.8%) |
|  | Administrative And Support And Waste Management And Remediation | 47,843 (8.1%) |
|  | Wholesale Trade | 33,598 (5.7%) |
|  | Utilities | 5,305 (0.9%) |
|  | Agriculture, Forestry, Fishing And Hunting | 5,248 (0.9%) |
|  | Mining, Quarrying, And Oil And Gas Extraction | 1,456 (0.2%) |
| Cause of Injury | Overexertion & bodily reaction | 155,499 (26.3%) |
|  | Struck by/against | 128,576 (21.7%) |
|  | Falls & slips | 116,350 (19.7%) |
|  | Motor vehicle | 48,349 (8.2%) |
|  | Contact with objects/equipment | 32,409 (5.5%) |
|  | Other/miscellaneous | 29,916 (5.1%) |
|  | Burns & exposure | 27,166 (4.6%) |
|  | Cut/puncture/scrape | 26,530 (4.5%) |
|  | Caught in/between | 15,877 (2.7%) |
|  | Assault & violence | 10,585 (1.8%) |
| Nature of Injury | Traumatic musculoskeletal | 398,811 (67.5%) |
|  | Open wounds & external trauma | 122,229 (20.7%) |
|  | Multiple / unspecified | 61,697 (10.4%) |
|  | Concussion / head injury | 4,972 (0.8%) |
|  | Systemic / internal conditions | 3,548 (0.6%) |
| Length of Employment | <3 months | 47,575 (8%) |
|  | 3-12 months | 66,393 (11.2%) |
|  | 1-2 years | 46,639 (7.9%) |
|  | ≥2 years | 221,791 (37.5%) |
|  | Missing | 208,859 (35.3%) |
| Time of Injury | Morning (6am-12pm) | 157,005 (26.6%) |
|  | Afternoon (12pm-6pm) | 142,470 (24.1%) |
|  | Evening/Night (6pm-6am) | 117,592 (19.9%) |
|  | Missing | 174,190 (29.5%) |
| Region | Downstate | 335,701 (56.8%) |
|  | Upstate | 255,556 (43.2%) |
| Benefit Type | Non-compensable | 341,198 (57.7%) |
|  | Medical Only | 59,180 (10%) |
|  | Temporary | 123,066 (20.8%) |
|  | Permanent Partial Disability (scheduled) | 60,754 (10.3%) |

|  |  |
| --- | --- |
| Permanent Partial Disability (non-scheduled) | 6,544 (1.1%) |
| Permanent Total Disability | 197 (0.0%) |
| Death | 318 (0.1%) |

**Notes:** Counts for age and gender may not sum to the total due to missing or unknown values. Downstate includes New York City counties plus Westchester, Nassau, and Suffolk. Non-compensable claims do not include indemnity payments but may include medical benefits if complete; these comprise both complete and incomplete claims. See Supplemental Table 4 for corresponding characteristics of complete and established claims.

Time of injury was coded to the nearest hour and grouped into morning (6:00 AM–11:59 AM), afternoon (12:00 PM–5:59 PM), and evening/night (6:00 PM–5:59 AM). County of injury was used to link meteorological exposures and define region. The downstate region included the five counties that comprise New York City, Westchester County, and Long Island (Nassau and Suffolk); all other counties were classified as upstate. Length of employment and time of injury were missing for a substantial proportion of claims and were retained as “missing” categories in stratified analyses.

### WC cost data

Claim cost data included Board-determined indemnity awards and insurer-reported paid benefits submitted through the electronic claims (eClaims) system, including medical, indemnity, and other benefit payments. When insurer-reported indemnity payment data were unavailable, Board-determined indemnity awards were used. Award amounts reflect compensation determined through formal Board decisions, whereas eClaims payment amounts represent totals reported by insurance carriers and are not independently validated by the Board.

### Weather Data

Daily meteorological data were obtained from the GridMET dataset (~4 km resolution) and spatially averaged to the county level.^33, 34^ We extracted daily maximum temperature, relative humidity, and total precipitation for NY during the study period and linked exposures to WC claims by county and date of injury.

Same-day maximum temperature was the primary exposure of interest, reflecting peak working conditions, alignment with policy-relevant thresholds, and avoiding limitations of heat index measures constructed from daily summary humidity values^35^. Mean daily heat index was evaluated in sensitivity analyses using the weathermetrics R package.^36^ Models were adjusted for daily precipitation (0 mm, 0–20 mm, >20 mm).

### Statistical Analysis

We used a time-stratified case-crossover design comparing heat exposure on the injury day with matched referent days, controlling for time-invariant characteristics and seasonality. Referent days were matched on year, month, day of week, and county of injury using a time-stratified approach; major U.S. holidays were excluded to avoid atypical work patterns.^37^

Conditional logistic regression models estimated associations between temperature and injury, with temperature modeled using a natural cubic spline (3 degrees of freedom), adjusted for precipitation, and 65°F as the reference. Effect modification by age, sex, industry sector, cause and nature of injury, length of employment, time of injury, and region was assessed using stratified models.

Primary dose–response models were estimated using assembled claims; identical models using complete and established claims assessed robustness to claim adjudication status. Robustness to heat metrics was evaluated by repeating models using heat index.

Attributable fractions (AFs) and 95% CIs were estimated for the entire warm season and for temperatures above policy-relevant thresholds (≥80°F and ≥90°F). AFs were derived from odds ratios (OR), which approximate incidence rate ratios (IRR) given the rarity of daily injury events, and calculated as AF = (IRR − 1)/IRR.

Annual excess injuries were estimated by applying AFs to mean annual warm-season claim counts. Excess injuries were estimated separately for complete and incomplete claims by applying AFs to subset-specific annual claim counts, distinguishing WC-captured from uncaptured costs potentially borne outside the WC system. Sensitivity analyses excluding pandemic years (2020–2021) yielded similar annual average claim counts; all years were retained.

### Cost estimation and cost-shifting

In the NYS WC system, only claims that progress to complete status contain payment data. Claims that remain incomplete retain a non-compensable designation and lack observed WC payments, although associated medical care may occur outside the WC system, shifting costs to private insurers, public programs, or workers.

WC-captured costs were estimated by applying a weighted mean total claim cost of $37,767 to the number of annual excess injuries attributable to heat. This value was derived from complete claims with nonzero payments in 2018–2019 (allowing ≥60 months of claim development), weighted by benefit type distribution among warm season claims (Supplemental Table 2). This estimate is consistent with the average WC claim cost reported by the New York Compensation Rating Board ($37,350 in 2022).^38^

Because costs for incomplete claims are not observed, we estimated a range. The lower bound applied mean medical benefit costs among zero-indemnity Medical Only claims ($3,389), representing a conservative estimate of medical expenditures. The upper bound applied the weighted mean total claim cost ($37,767), representing a plausible upper limit if incomplete claims were similar in severity to complete claims.

Costs are reported separately for complete (WC-captured) and incomplete claims.

This study followed STROBE reporting guidelines for case-control studies, consistent with the case-crossover design.

## Results

### I. Claim descriptives

Among 591,257 assembled claims from 2016–2024, claimants were predominantly male (73.9%), and 39.5% were aged 35 years or younger. Public administration (e.g., municipal and state employees, police, inspectors, sanitation, and maintenance staff) accounted for the largest share of claims (30.3%), followed by transportation and warehousing (15.9%), manufacturing (13.9%), accommodation and food services (13.2%), and construction (10.8%) (Table 1).

Overexertion (26.3%), struck-by incidents (21.7%), and falls (19.7%) accounted for most injuries. By nature of injury, traumatic musculoskeletal conditions predominated (67.5%), primarily strains, sprains, contusions, and fractures. Systemic and internal conditions were rare (0.6%) but included the most directly coded heat-related illnesses (e.g., heat prostration; n=1,325) (Supplemental Table 3).

Overall, 71.0% of assembled claims progressed to complete status and 42.3% to established status; most were non-compensable, medical-only, or temporary disability (88.5%) (Supplemental Tables 4).

### II. Weather descriptives

Each case was matched to an average of 3.34 referent days. Distributions of temperature, heat index, humidity, and precipitation were highly similar between case and referent days, indicating effective control for time-invariant and seasonal factors (Supplemental Table 5).

### III. Heat-injury odds ratios and effect modification

Occupational injury risk increased steadily with higher temperatures (Figure 2; Supplemental Table 6). Relative to 65°F, injury odds increased to 1.06 (95% CI: 1.01–1.10) at 80°F, 1.12 (1.07–1.18) at 90°F, and 1.17 (1.11–1.23) at 95°F (Figure 2, Panel A). Associations became statistically significant at approximately 78°F.

**Figure 2:**
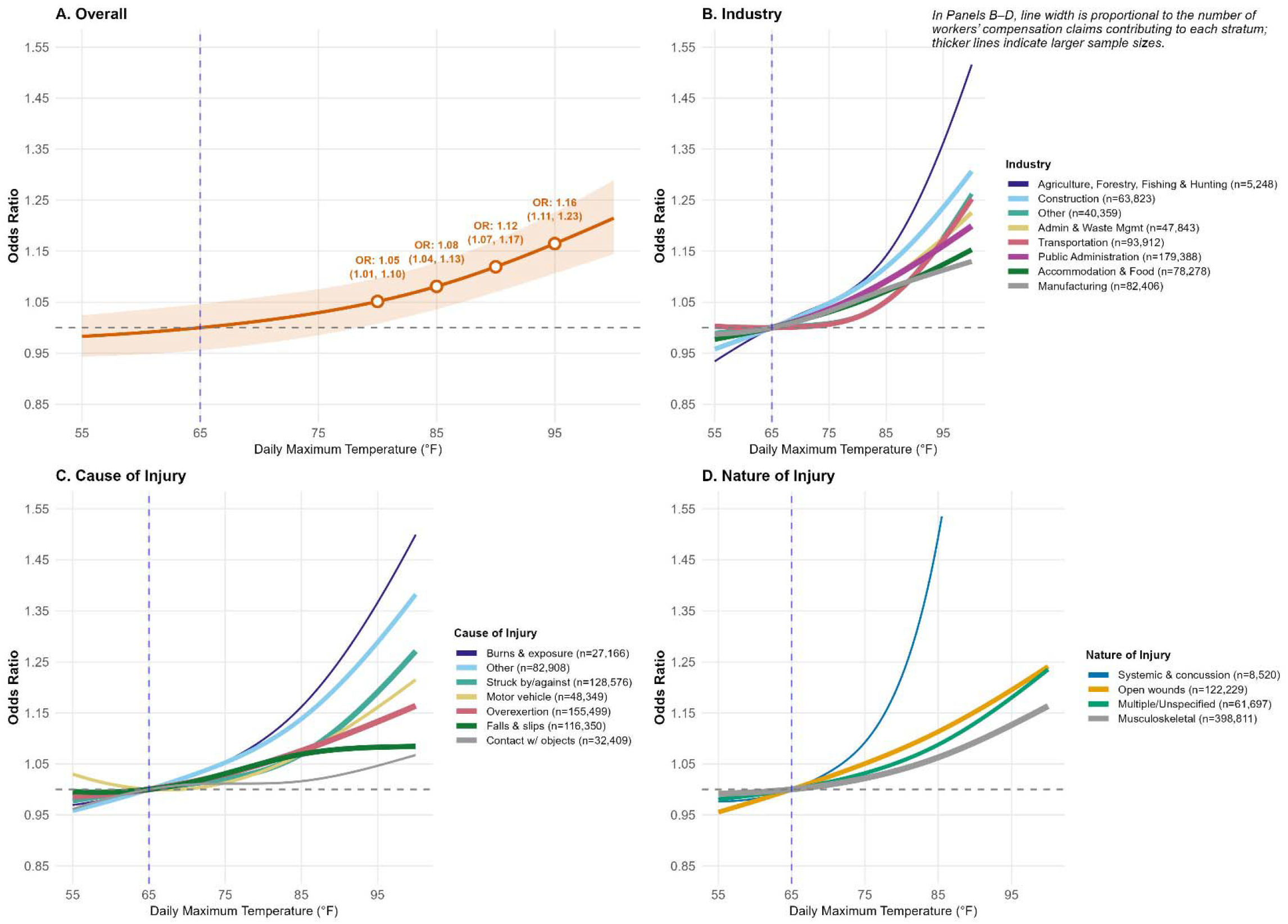
Heat-injury dose-response relationship for assembled New York workers’ compensation claims overall and by industry, and cause and nature of injury. **Notes:** Odds ratios were estimated using time-stratified case-crossover models with conditional logistic regression and natural cubic splines (3 degrees of freedom), adjusted for precipitation. Estimates and 95% CIs at selected temperatures are reported in Supplemental Table 6. Some categories were collapsed due to small sample sizes. “Other industry” includes utilities, wholesale trade, and administrative and support and waste management and remediation services. “Other/miscellaneous” cause includes infrequent categories not shown separately. Systemic/internal conditions and concussion/head injury were combined.

Results were consistent across assembled, complete, and established claims with nearly identical point estimates and were robust to use of heat index (Supplemental Figures 1–2).

Industry-stratified analyses showed similar upward temperature-injury trends but differing magnitudes across sectors (Figure 2, Panel B). Injury risk became statistically significant at 85°F in public administration (OR 1.10, 95% CI: 1.02–1.19), 90°F in construction (1.18, 1.02–1.37), and 95°F in transportation and warehousing (OR 1.19, 1.03–1.37). Agriculture showed the highest point estimates but with wide CIs (Supplemental Table 6).

By cause of injury, modest but significant increases in risk were observed in large claim groups, including overexertion and struck-by/against injuries (OR 1.13 95% CI 1.03–1.25 and OR 1.20 95%CI 1.07–1.34 at 95°F; Figure 2, Panel C; Supplemental Table 5). Smaller categories, including burns and environmental exposures and other/miscellaneous causes, showed steeper increases (ORs between 1.37 - 1.48 at 95°F). In contrast, falls, motor vehicle, and contact-related injuries showed weaker associations overall; motor vehicle injuries exhibited a modest upward trend, but confidence intervals crossed the null, while falls showed relatively flat temperature–risk patterns.

By nature of injury, traumatic musculoskeletal and open-wound injuries, which account for the majority of claims, increased gradually with temperature, reaching 13%–19% higher odds at 95°F versus 65°F (Figure 2, Panel D; Supplemental Table 5). The combined systemic/internal and concussion category exhibited a steep, nonlinear increase in injury odds beginning around 75–80°F. When examined separately, this pattern was driven by systemic conditions (e.g., heat prostration, myocardial infarction), which were associated with more than a tenfold increase in odds at 95°F (OR 10.17, 95% CI 4.93–20.98), rather than concussion diagnoses.

Associations were generally stronger among male, younger (ages 16–35), and shorter-tenure workers, although estimates were imprecise in some subgroups. Elevated odds were also observed among workers with missing job tenure and time of injury beginning at ≥85°F. Patterns were similar across upstate and downstate regions (Supplemental Figures 3-7).

### IV. Attributable fraction and economic burden

Heat exposure accounted for a sizable burden of occupational injury and costs (Table 2). Among complete claims, heat exposure during the warm season resulted in 2,322 excess claims annually (AF: 5.0%, 95% CI: 0.7– 9.1), corresponding to $87.7 million ($12.6–$159.5 million) in WC-captured costs. At ≥80°F, this included 1,221 excess claims and $46.1 million in WC costs. Although the AF was highest at ≥90°F (12.2%), these days were rare and accounted for fewer excess claims (194 annually; $7.3 million) (Table 2: Panel A).

**Table 2:**
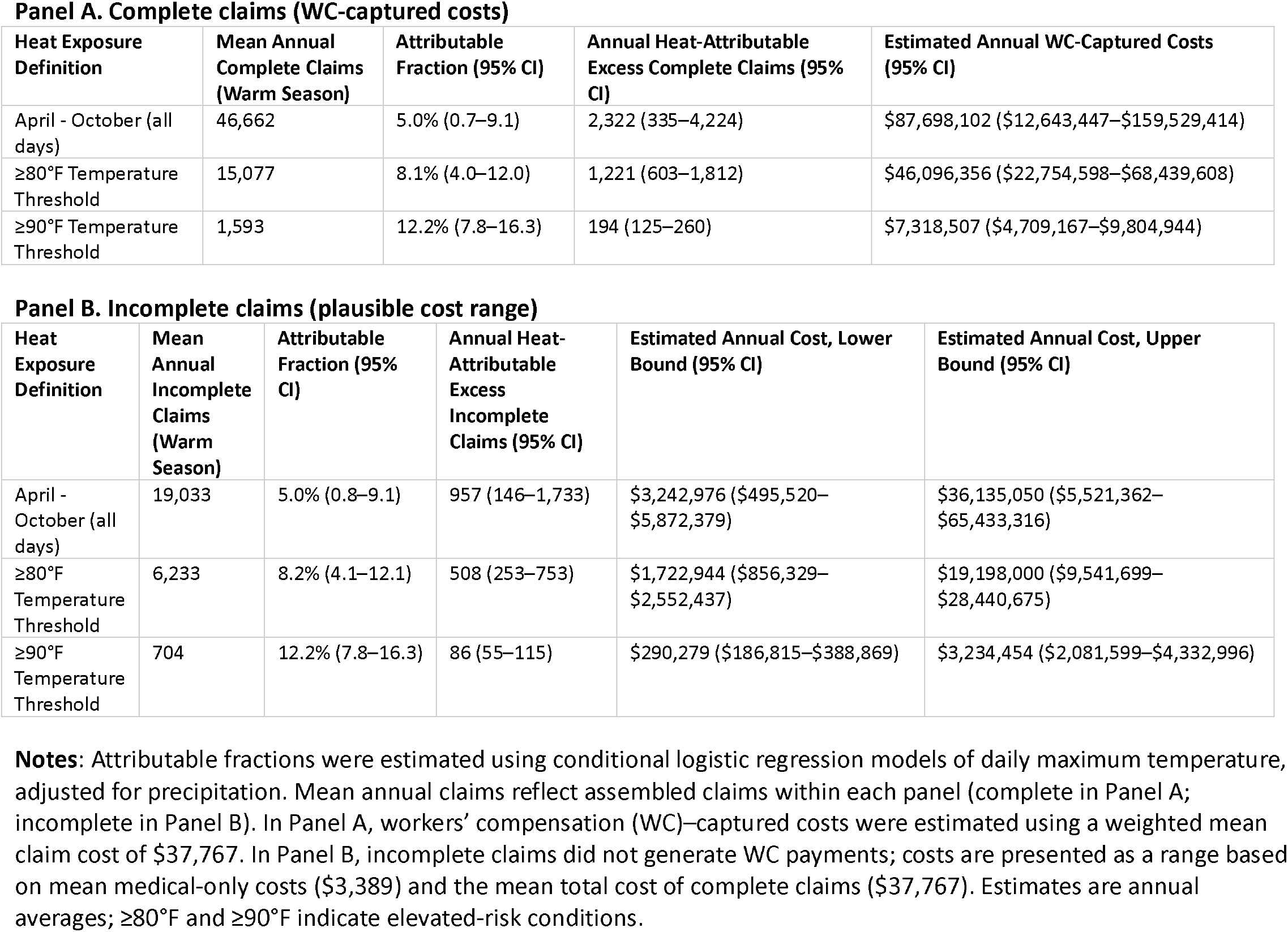
Estimated annual heat-attributable workers’ compensation claims and costs in New York State, by claim completeness, warm season (April–October), 2016–2024.

**Panel A. Complete claims (WC-captured costs)**
| Heat Exposure Definition | Mean Annual Complete Claims (Warm Season) | Attributable Fraction (95% CI) | Annual Heat-Attributable Excess Complete Claims (95% CI) | Estimated Annual WC-Captured Costs (95% CI) |
| --- | --- | --- | --- | --- |
| April - October (all days) | 46,662 | 5.0% (0.7–9.1) | 2,322 (335–4,224) | \$87,698,102 (\$12,643,447–\$159,529,414) |
| ≥80°F Temperature Threshold | 15,077 | 8.1% (4.0–12.0) | 1,221 (603–1,812) | \$46,096,356 (\$22,754,598–\$68,439,608) |
| ≥90°F Temperature Threshold | 1,593 | 12.2% (7.8–16.3) | 194 (125–260) | \$7,318,507 (\$4,709,167–\$9,804,944) |

**Panel B. Incomplete claims (plausible cost range)**
| Heat Exposure Definition | Mean Annual Incomplete Claims (Warm Season) | Attributable Fraction (95% CI) | Annual Heat-Attributable Excess Incomplete Claims (95% CI) | Estimated Annual Cost, Lower Bound (95% CI) | Estimated Annual Cost, Upper Bound (95% CI) |
| --- | --- | --- | --- | --- | --- |
| April - October (all days) | 19,033 | 5.0% (0.8–9.1) | 957 (146–1,733) | \$3,242,976 (\$495,520–\$5,872,379) | \$36,135,050 (\$5,521,362–\$65,433,316) |
| ≥80°F Temperature Threshold | 6,233 | 8.2% (4.1–12.1) | 508 (253–753) | \$1,722,944 (\$856,329–\$2,552,437) | \$19,198,000 (\$9,541,699–\$28,440,675) |
| ≥90°F Temperature Threshold | 704 | 12.2% (7.8–16.3) | 86 (55–115) | \$290,279 (\$186,815–\$388,869) | \$3,234,454 (\$2,081,599–\$4,332,996) |

**Notes:** Attributable fractions were estimated using conditional logistic regression models of daily maximum temperature, adjusted for precipitation. Mean annual claims reflect assembled claims within each panel (complete in Panel A; incomplete in Panel B). In Panel A, workers' compensation (WC)–captured costs were estimated using a weighted mean claim cost of \$37,767. In Panel B, incomplete claims did not generate WC payments; costs are presented as a range based on mean medical-only costs (\$3,389) and the mean total cost of complete claims (\$37,767). Estimates are annual averages; ≥80°F and ≥90°F indicate elevated-risk conditions.

A meaningful share of this burden occurred among incomplete claims, with costs likely borne outside the WC system (Table 2: Panel B). We estimated 957 excess incomplete claims annually (95% CI: 146–1,733). At ≥80°F, this corresponded to 508 excess claims (95% CI: 253–753), with costs ranging from $1.7 million to $19.2 million.

## Discussion

Occupational injury risk increased steadily with rising ambient temperatures in NY, with a clear dose– response relationship and statistically significant effects beginning at approximately 78°F. The magnitude of risk at common temperature thresholds was consistent with prior studies from warmer U.S. settings, demonstrating that similar heat–injury relationships occur in a large Northeastern labor market where temperatures are cooler.^12, 14, 16^ These findings strengthen the case for national workplace heat protections and highlight that harmful effects occur well below extreme heat thresholds.

Heat-attributable occupational injuries, most of which are not explicitly coded as heat-related, impose substantial and largely preventable economic costs. At temperatures ≥80°F, we estimate approximately 1,729 excess injuries annually—about five per day—generating $46.4 million in WC expenditures. In contrast, injuries explicitly coded as heat-related amounted to less than $1 million annually over the study period (data not shown), underscoring the extent to which traditional surveillance underestimates heat-related occupational burden.

An 80°F threshold aligns with proposed policy temperature thresholds, indicating that many injuries occur under preventable conditions. Although extreme heat (≥90°F) produced the highest relative risks, such days were rare in NY; most heat-attributable injuries occurred on more common warm days, highlighting the importance of early intervention. Under projected warming of approximately 3.6°F in NY by 2050, this burden is likely to increase substantially.^39^

Evidence from jurisdictions with heat standards suggests that prevention strategies can reduce heat-related harm. In California, which implemented the first comprehensive heat illness prevention standard for outdoor workers in 2005, heat-related deaths among outdoor workers declined relative to neighboring states without such standards.^40^ Dean and McCallum (2025) found a 33% decline in fatalities following enhanced enforcement (2010–2014) and a 51% decline after implementation of a revised standard (2015–2020).^40^ Analyses of California WC claims similarly show declines in injury filings on hot days across heat-exposed industries after implementation of the standard.^14^ A recent nationwide analysis also found evidence of attenuated injury risk on hot days in states with occupational heat standards.^16^

Prior research shows that many work-related injuries are never reported or compensated, shifting medical costs to private insurance, public programs, or workers themselves.^41-44^ Our findings extend this literature by suggesting that similar cost shifting may occur even among injuries that are reported but do not progress through the WC system. We estimate approximately 957 heat-attributable incomplete claims annually, with associated costs of $3.2-$36.1 million.

These uncompensated injuries include traumatic musculoskeletal conditions and selected systemic illnesses that can generate substantial medical expenditures but may be difficult to adjudicate as compensable. Notably, they were reported to employers and entered the WC system, indicating that workers had already overcome key barriers to filing a claim. These findings suggest that WC costs understate the economic burden of occupational heat exposure. Further research is needed to understand why many claims do not progress through the WC system and are not captured in standard reporting, including the NYS WCB annual report.^45^

WC expenditures capture only part of the economic burden of occupational heat exposure. Heat also reduces labor capacity and productivity.^46^ One analysis estimated that extreme heat results in the loss of approximately 421 million work hours annually in the U.S., equivalent to about $14 billion.^47^ These losses exceed the estimated national compliance costs of the proposed federal heat standard (approximately $7.8 billion annually).^1^ Similarly, an analysis of California’s proposed indoor heat standard estimated annual compliance costs of $102 million compared with benefits of approximately $402 million.^48^

Effect-modification findings further inform prevention strategies. Elevated risks across diverse industries and among newer workers underscore the importance of acclimatization and graduated exposure protocols. Increased risks in both downstate and cooler upstate regions indicate that protections should not be limited to traditionally high-temperature geographies. Finally, the sharp increase in systemic heat illness alongside more gradual increases in traumatic injuries suggests that prevention training must address both direct heat illness and indirect pathways, including fatigue, reduced attention, and impaired decision-making.

This study has several limitations. WC data lack information on race and ethnicity, limiting assessment of disparities in heat-related injury risk and access to benefits. This is particularly important given evidence that low-wage and immigrant workers, who are disproportionately represented in high heat–exposure industries, face both elevated occupational risks and structural barriers to reporting and claim filing.^4, 6, 18, 49, 50^ As a result, WC data likely undercapture the true burden of occupational injury, with the greatest underrepresentation among populations most affected by heat, potentially biasing estimates downward. Incomplete claims may also reflect barriers to claim progression that disproportionately affect marginalized workers; however, these mechanisms could not be directly assessed in our data.

Additional limitations include cost estimates based on paid amounts in administrative data, which may not reflect total medical expenditures; we were unable to identify payers for care associated with incomplete claims. Temperature exposure was based on ambient outdoor conditions and may not reflect individual exposure due to work schedules, indoor environments, workload, protective equipment, or microclimates; daily measures may also not align precisely with working hours. Future research should examine barriers to claim progression and whether administrative reforms could reduce cost-shifting and improve equitable access to benefits.

## Conclusion

As temperatures rise, occupational heat exposure will increasingly contribute to preventable injury, medical spending, and productivity loss. Much of this burden occurs during routine warm conditions and extends beyond injuries explicitly coded as heat-related or costs captured in the WC system. Despite growing evidence of harm, workplace heat protections remain limited in the U.S. Heat prevention should therefore be viewed not only as an occupational safety priority but also as a cost-containment strategy that can reduce health care expenditures and strengthen workforce resilience in a warming climate.

## Data Availability

All data used in this study were obtained from publicly available sources, including New York State Workers' Compensation Board claims data and publicly available meteorological datasets. No new data were generated.

https://data.ny.gov/Government-Finance/Assembled-Workers-Compensation-Claims-Beginning-20/jshw-gkgu/about_data

https://www.climatologylab.org/gridmet.html

## Acknowledgments

We thank June Spector, Les Boden, and the New York Compensation Insurance Rating Board for their time and insight in developing the methods for this paper. We also thank the New York State Workers’ Compensation Board for clarifying administrative questions.

