## Supplemental Materials for "Heat Exposure, Occupational Injury Risk, and Economic Costs in New York State"

### Supplemental Methods I: Rationale for using Assembled claims as primary analytic sample

To identify the workers’ compensation (WC) claim subset that best captures population-level occupational injury incidence, we compared annual counts from New York State Workers’ Compensation Board (NYS WCB) claims with employer-reported occupational injuries and fatalities from the Bureau of Labor Statistics (BLS) Survey of Occupational Injuries and Illnesses (SOII) and the Census of Fatal Occupational Injuries (CFOI) for all available years. BLS data for 2024 were not yet available and were therefore excluded from these comparisons (Supplemental Table 1).

Because employer-based injury surveillance is known to undercount occupational injuries, SOII estimates were adjusted upward using 20% and 60% correction factors reflecting published estimates of underreporting. After applying these adjustments, annual counts of assembled WC claims consistently fell within the range of corrected BLS estimates, suggesting that assembled claims provide the most complete representation of occupational injury incidence in New York State.

In contrast, fewer claims progressed to complete or established status. Although reasons for non-progression are not observed in the data, prior research suggests that administrative and structural barriers can limit claim progression. Restricting analyses to these subsets would preferentially capture more severe or successfully adjudicated cases and introduce selection bias. Accordingly, assembled claims were used for primary analyses, while complete and established claims were used for sensitivity and cost analyses, where payment data are required. Because incomplete claims lack observed payment data, their associated costs were estimated separately using a range of plausible values.

### Supplemental Table 1: Comparison of occupational injuries and fatalities in New York State: BLS surveillance and workers' compensation claims data, 2016-2024

| **Accident Year** | **BLS SOII** | **BLS CFOI** | **BLS Total (SOII + CFOI)** | **BLS Total (Adjusted for 20% Underreporting)** | **Total (Adjusted for 60% Underreporting)** | **NYS WC-ASSEMBLED** | **NYS WC - COMPLETE** | **NYS WC– ESTABLISHED** |
| --- | --- | --- | --- | --- | --- | --- | --- | --- |
| 2016 | 200,500 | 272 | 200,772 | 240,872 | 321072 | 266607 | 168217 | 94256 |
| 2017 | 203,100 | 313 | 203,413 | 244,033 | 325273 | 275482 | 175479 | 96236 |
| 2018 | 200,600 | 271 | 200,871 | 240,991 | 321231 | 288579 | 183622 | 100758 |
| 2019 | 206,700 | 278 | 206,978 | 248,313 | 330993 | 291545 | 182539 | 101072 |
| 2020 | 189,300 | 223 | 189,523 | 227,383 | 303103 | 224808 | 134205 | 82265 |
| 2021 | 183,700 | 247 | 183,947 | 220,687 | 294167 | 246608 | 146064 | 91558 |
| 2022 | 192,800 | 251 | 193,051 | 231,611 | 308731 | 262107 | 158701 | 93556 |
| 2023 | 193,200 | 246 | 193,446 | 232,086 | 309366 | 259735 | 163279 | 89282 |
| 2024 | -- | -- | -- | -- | -- | 258362 | 158212 | 75393 |
| **TOTAL** | **1,569,900** | **2,096** | **1,571,996** | **1,885,976** | **2,513,936** | **2,373,833** | **1,470,318** | **824,376** |

**Source:** Annual occupational injury and fatality counts were obtained from the Bureau of Labor Statistics (BLS) Survey of Occupational Injuries and Illnesses (SOII) and Census of Fatal Occupational Injuries (CFOI). New York State (NYS) Workers’ Compensation (WC) claims data were analyzed by the authors using publicly available data from data.ny.gov.

**Notes:** SOII captures nonfatal injuries and illnesses meeting reporting criteria; CFOI captures all fatal occupational injuries. BLS totals were adjusted upward by 20% and 60% to account for underreporting. Assembled claims include all reported cases with a Board file; complete claims include insurer notice and medical documentation; established claims include a determination of work-relatedness. BLS data for 2024 were unavailable. NYS WC does not cover some workers (e.g., federal employees, certain public employees, independent contractors, and gig workers).

### Supplemental Table 2: Weighted mean claim cost calculation using 2018 - 2019 benefit type distribution on warm days (≥80°F)

| **Benefit Type** | **No. of Claims (Apr -Oct, 2018-2019)** | **Percent** | **Mean Total Cost ($)** | **Weighted Mean Claim Cost ($)** |
| --- | --- | --- | --- | --- |
| Medical Only / Non-compensable | 72713 | 53.59 | 5948.95 | 3187.92 |
| Temporary | 40460 | 29.82 | 66370.40 | 19790.45 |
| Permanent Partial Disability (scheduled) | 20126 | 14.83 | 70486.19 | 10454.83 |
| Permanent Partial Disability (non-scheduled) | 2229 | 1.64 | 233934.73 | 3842.91 |
| Death | 100 | 0.07 | 201395.17 | 148.42 |
| Permanent Total Disability | 61 | 0.04 | 761317.07 | 342.26 |
| **Weighted mean total claim cost** |  |  |  | **37,766.79** |

**Source:** New York State Workers’ Compensation Board (NYS WCB) claims data, including insurer-reported benefit payments submitted through the electronic claims (eClaims) system and Board-determined indemnity awards.

**Notes:** Total claim costs include medical, indemnity, and other benefit payments. Costs were calculated using complete and established claims with nonzero payments occurring on warm days (daily maximum temperature ≥80°F) between April and October in 2018–2019. Mean total costs were estimated by benefit type and combined using the observed warm-day benefit-type distribution to derive the weighted mean WC claim cost. Medical-only and complete non-compensable claims were combined because both lack indemnity payments; non-compensable claims may not be formally established as compensable, yet medical payments can still occur.

### Supplemental Table 3. Detailed cause and nature of injury for all assembled claims in analytic sample, (n=591,257), NYS, 2016-2024

| **Type** | **Broad Category** | **Specific Descriptions** | **No. of Claims** |
| --- | --- | --- | --- |
| **Cause of Injury** | | | |
|  | **Assault & violence** | |  |
|  |  | Person in act of a crime | 10,484 |
|  |  | Gunshot | 101 |
|  | **Burns & exposure** | |  |
|  |  | Foreign matter (body) in eye(s) | 7,914 |
|  |  | Hot objects or substances | 5,929 |
|  |  | Steam or hot fluids | 3,678 |
|  |  | Chemicals | 2,592 |
|  |  | Absorption, ingestion or inhalation, NOC | 2,367 |
|  |  | Electrical current | 1,204 |
|  |  | Temperature extremes | 1,034 |
|  |  | Dust, gases, fumes or vapors | 1,021 |
|  |  | Fire or flame | 961 |
|  |  | Welding operation | 228 |
|  |  | Explosion or flare back | 218 |
|  |  | Mold | 20 |
|  | **Caught in/between** | |  |
|  |  | Object handled | 11,223 |
|  |  | Caught in, under or between, NOC | 9,328 |
|  |  | Machine or machinery | 6,549 |
|  | **Contact with objects/equipment** | |  |
|  |  | Hand tool, utensil; not powered | 14,133 |
|  |  | Powered hand tool, appliance | 6,136 |
|  |  | Hand tool or machine in use | 5,141 |
|  |  | Using tool or machinery | 4,262 |
|  |  | Moving parts of machine | 1,948 |
|  |  | Moving part of machine | 789 |
|  | **Cut/puncture/scrape** | |  |
|  |  | Cut, puncture, scrape, NOC | 18,570 |
|  |  | Broken glass | 5,686 |
|  |  | Stepping on sharp object | 2,274 |
|  | **Falls & slips** |  |  |
|  |  | Fall, slip or trip, NOC | 32,305 |
|  |  | On same level | 28,483 |
|  |  | From different level (elevation) | 15,469 |
|  |  | On stairs | 12,961 |
|  |  | From liquid or grease spills | 9,606 |
|  |  | From ladder or scaffolding | 9,177 |
|  |  | Slip, or trip, did not fall | 5,680 |
|  |  | Into openings | 2,669 |
|  | **Motor vehicle** |  |  |
|  |  | Collision or sideswipe with another vehicle | 32,483 |
|  |  | Motor vehicle, NOC | 6,889 |
|  |  | Motor vehicle | 5,891 |
|  |  | Collision with a fixed object | 2,011 |
|  |  | Vehicle upset | 956 |
|  |  | Crash of rail vehicle | 69 |
|  |  | Crash of water vehicle | 39 |
|  |  | Crash of airplane | 11 |
|  | **Other/miscellaneous** | |  |
|  |  | Other - miscellaneous, NOC | 24,199 |
|  |  | Contact with, NOC | 5,128 |
|  |  | Rubbed or abraded, NOC | 346 |
|  |  | Sanding, scraping, cleaning operation | 219 |
|  |  | Natural disasters | 24 |
|  | **Overexertion & bodily reaction** | |  |
|  |  | Lifting | 51,871 |
|  |  | Strain or injury by, NOC | 38,508 |
|  |  | Pushing or pulling | 32,066 |
|  |  | Twisting | 14,619 |
|  |  | Holding or carrying | 8,356 |
|  |  | Reaching | 5,169 |
|  |  | Cumulative, NOC | 2,627 |
|  |  | Jumping or leaping | 1,802 |
|  |  | Wielding or throwing | 481 |
|  | **Struck by/against** | |  |
|  |  | Object being lifted or handled | 36,008 |
|  |  | Falling or flying object | 24,080 |
|  |  | Fellow worker, patient or other person | 17,216 |
|  |  | Struck or injured, NOC | 15,972 |
|  |  | Stationary object | 15,018 |
|  |  | Striking against or stepping on, NOC | 5,445 |
|  |  | Object handled by others | 3,614 |
| **Nature of Injury** | |  |  |
|  | **Concussion / head injury** | |  |
|  |  | Concussion | 4,972 |
|  | **Multiple / unspecified** | |  |
|  |  | All other specific injuries, NOC | 46,896 |
|  |  | Multiple physical injuries only | 14,428 |
|  |  | Multiple injuries including both physical and psychological | 373 |
|  | **Open wounds & external trauma** | |  |
|  |  | Laceration | 78,132 |
|  |  | Burn | 14,504 |
|  |  | Foreign body | 13,368 |
|  |  | Puncture | 12,025 |
|  |  | Dermatitis | 1,635 |
|  |  | Amputation | 1,080 |
|  |  | Electric shock | 1,065 |
|  |  | Severance | 420 |
|  | **Systemic / internal conditions** | |  |
|  |  | Syncope | 1,387 |
|  |  | Heat prostration | 1,325 |
|  |  | Myocardial infarction | 518 |
|  |  | Angina pectoris | 318 |
|  | **Traumatic musculoskeletal** | |  |
|  |  | Strain or tear | 172,098 |
|  |  | Contusion | 114,216 |
|  |  | Sprain or tear | 72,449 |
|  |  | Fracture | 25,430 |
|  |  | Crushing | 7,230 |
|  |  | Dislocation | 6,213 |
|  |  | Rupture | 1,175 |

**Source:** Authors’ analysis of New York State Workers’ Compensation Board (NYS WCB) claims data, 2016–2024.

**Notes:** Cause and nature of injury were classified using the Workers’ Compensation Insurance Organizations (WCIO) Injury Description Codes for cause of injury and nature of injury, as reported in NYS WCB administrative records. Broad analytic categories group detailed WCIO codes reflecting similar injury mechanisms (cause) or clinical outcomes (nature). NOC denotes “not otherwise classified” WCIO codes.

### Supplemental Table 4: Detailed descriptive statistics of assembled, complete, and established NYS Workers' Compensation Claims

| **Variable** | **Level** | **Assembled (N=591,257)** | **Complete (N=419,956)** | **Established (N=250,059)** |
| --- | --- | --- | --- | --- |
| **Age Category (years)** | 16-25 | 80,441 (13.6%) | 50,142 (12.0%) | 23,454 (9.4%) |
|  | 26-35 | 152,436 (25.8%) | 105,482 (25.2%) | 61,445 (24.6%) |
|  | 36-45 | 132,799 (22.5%) | 97,213 (23.2%) | 61,449 (24.6%) |
|  | 46-55 | 126,738 (21.5%) | 94,061 (22.4%) | 59,542 (23.8%) |
|  | 56+ | 97,649 (16.5%) | 72,336 (17.3%) | 43,794 (17.5%) |
| **Gender** | Female | 152,884 (26.1%) | 102,120 (24.5%) | 59,310 (23.8%) |
|  | Male | 433,645 (73.9%) | 315,161 (75.5%) | 189,494 (76.2%) |
| **Industry** | Public Administration | 179,388 (30.3%) | 127,556 (30.4%) | 82,435 (33.0%) |
|  | Transportation And Warehousing | 93,912 (15.9%) | 71,778 (17.1%) | 46,354 (18.5%) |
|  | Manufacturing | 82,406 (13.9%) | 59,458 (14.2%) | 29,193 (11.7%) |
|  | Accommodation And Food Services | 78,278 (13.2%) | 46,059 (11.0%) | 22,909 (9.2%) |
|  | Construction | 63,823 (10.8%) | 48,464 (11.5%) | 31,761 (12.7%) |
|  | Administrative And Support And Waste Management And Remediation | 47,843 (8.1%) | 33,191 (7.9%) | 18,879 (7.5%) |
|  | Wholesale Trade | 33,598 (5.7%) | 24,248 (5.8%) | 13,501 (5.4%) |
|  | Utilities | 5,305 (0.9%) | 4,171 (1.0%) | 2,228 (0.9%) |
|  | Agriculture, Forestry, Fishing And Hunting | 5,248 (0.9%) | 3,990 (1.0%) | 2,242 (0.9%) |
|  | Mining, Quarrying, And Oil And Gas Extraction | 1,456 (0.2%) | 1,041 (0.2%) | 557 (0.2%) |
| **Cause of Injury** | Overexertion & bodily reaction | 155,499 (26.3%) | 118,536 (28.2%) | 76,149 (30.5%) |
|  | Struck by/against | 128,576 (21.7%) | 84,084 (20.0%) | 45,065 (18.0%) |
|  | Falls & slips | 116,350 (19.7%) | 87,477 (20.8%) | 58,050 (23.2%) |
|  | Motor vehicle | 48,349 (8.2%) | 40,483 (9.6%) | 27,478 (11.0%) |
|  | Contact with objects/equipment | 32,409 (5.5%) | 21,422 (5.1%) | 9,921 (4.0%) |
|  | Other/miscellaneous | 29,916 (5.1%) | 18,536 (4.4%) | 10,201 (4.1%) |
|  | Burns & exposure | 27,166 (4.6%) | 14,372 (3.4%) | 5,022 (2.0%) |
|  | Cut/puncture/scrape | 26,530 (4.5%) | 14,991 (3.6%) | 5,017 (2.0%) |
|  | Caught in/between | 15,877 (2.7%) | 11,899 (2.8%) | 6,985 (2.8%) |
|  | Assault & violence | 10,585 (1.8%) | 8,156 (1.9%) | 6,171 (2.5%) |
| **Nature of Injury** | Traumatic musculoskeletal | 398,811 (67.5%) | 301,683 (71.8%) | 195,761 (78.3%) |
|  | Open wounds & external trauma | 122,229 (20.7%) | 71,407 (17.0%) | 27,052 (10.8%) |
|  | Multiple / unspecified | 61,697 (10.4%) | 41,411 (9.9%) | 24,478 (9.8%) |
|  | Concussion / head injury | 4,972 (0.8%) | 3,736 (0.9%) | 2,281 (0.9%) |
|  | Systemic / internal conditions | 3,548 (0.6%) | 1,719 (0.4%) | 487 (0.2%) |
| **Length of Employment** | <3 months | 47,575 (8%) | 31,025 (7.4%) | 16,029 (6.4%) |
|  | 3-12 months | 66,393 (11.2%) | 43,306 (10.3%) | 22,666 (9.1%) |
|  | 1-2 years | 46,639 (7.9%) | 31,305 (7.5%) | 16,797 (6.7%) |
|  | ≥2 years | 221,791 (37.5%) | 160,549 (38.2%) | 97,137 (38.8%) |
|  | Missing | 208,859 (35.3%) | 153,771 (36.6%) | 97,430 (39%) |
| **Time of Injury** | Morning (6am-12pm) | 157,005 (26.6%) | 111,342 (26.5%) | 62,666 (25.1%) |
|  | Afternoon (12pm-6pm) | 142,470 (24.1%) | 98,703 (23.5%) | 56,201 (22.5%) |
|  | Evening/Night (6pm-6am) | 117,592 (19.9%) | 82,923 (19.7%) | 48,573 (19.4%) |
|  | Missing | 174,190 (29.5%) | 126,988 (30.2%) | 82,619 (33.0%) |
| **Region** | Downstate | 335,701 (56.8%) | 237,305 (56.5%) | 154,390 (61.7%) |
|  | Upstate | 255,556 (43.2%) | 182,651 (43.5%) | 95,669 (38.3%) |
| **Benefit Type** | Non-compensable | 341,198 (57.7%) | 169,897 (40.5%) |  |
|  | Medical Only | 59,180 (10.0%) | 59,180 (14.1%) | 59,180 (23.7%) |
|  | Temporary | 123,066 (20.8%) | 123,066 (29.3%) | 123,066 (49.2%) |
|  | Permanent Partial Disability (scheduled) | 60,754 (10.3%) | 60,754 (14.5%) | 60,754 (24.3%) |
|  | Permanent Partial Disability (non-scheduled) | 6,544 (1.1%) | 6,544 (1.6%) | 6,544 (2.6%) |
|  | Permanent Total Disability | 197 (0.0%) | 197 (0.0%) | 197 (0.1%) |
|  | Death | 318 (0.1%) | 318 (0.1%) | 318 (0.1%) |

**Source:** Authors’ analysis of New York State Workers’ Compensation Board claims data, 2016–2024.

**Notes**: Counts for age and gender may not sum to the total due to missing or unknown values. Downstate includes New York City counties plus Westchester, Nassau, and Suffolk. Non-compensable claims did not result in indemnity awards but may include paid medical benefits if they progressed to complete status.

### Supplemental Table 5. Distribution of weather variables for case and referent days

| **Weather Variables** | **Referent Days (n=1,975,074)** | **Case Days (n=591,257)** |
| --- | --- | --- |
|  | Max (Median, Interquartile Range) | Max (Median, Interquartile Range) |
| Max Temperature (°F) | 98.5 (75.1, 16.1) | 98.5 (75.2, 16.1) |
| Mean Heat Index (°F) | 64.1 (66.0, 17.0) | 64.3 (66.0, 17.0) |
| Mean Relative Humidity (%) | 68.0 (69.3, 13.9) | 68.1 (69.4, 13.8) |
| Precipitation (mm) | 178.5 (0.0, 3.1) | 178.5 (0.0, 3.1) |
| Case to Referent Day Ratio = 1:3.34 |  |  |

**Source:** Authors’ analysis of New York State Workers’ Compensation Board claims, 2016–2024, and GridMET meteorological data.

**Notes:** Case days correspond to injury dates; referent days were selected using a time-stratified case-crossover design matched on year, month, day of week, and county. Similar distributions between case and referent days reflect appropriate matching and control for temporal confounding.

### Supplemental Table 6: Odds ratios and 95% confidence intervals for max daily temperature and injury relationship at 80, 85, 90 and 95 degrees

|  |  | **OR (95% CI)** | **OR (95% CI)** | **OR (95% CI)** | **OR (95% CI)** |
| --- | --- | --- | --- | --- | --- |
| Variable | **Category (No. of claims)** | 80°F | 85°F | 90°F | 95°F |
| **Overall** | Overall (n=591,257) | 1.057 (1.013-1.102) | 1.087 (1.041-1.134) | 1.123 (1.072-1.176) | 1.165 (1.106-1.227) |
| Age (years) | 16-25 (n= 80,441) | 1.082 (0.964-1.214) | 1.123 (0.999-1.262) | 1.172 (1.034-1.329) | 1.228 (1.066-1.414) |
|  | 26-35 (n=152,436) | 1.057 (0.972-1.150) | 1.090 (1.001-1.187) | 1.131 (1.032-1.240) | 1.179 (1.064-1.306) |
|  | 36-45 (n=132,799) | 1.045 (0.954-1.145) | 1.073 (0.978-1.178) | 1.112 (1.006-1.228) | 1.157 (1.035-1.293) |
|  | 46-55 (n=126,738) | 1.044 (0.955-1.141) | 1.068 (0.976-1.170) | 1.099 (0.997-1.211) | 1.134 (1.016-1.265) |
|  | 56+ (n= 97,649) | 1.066 (0.967-1.174) | 1.091 (0.989-1.205) | 1.116 (1.003-1.241) | 1.139 (1.010-1.286) |
| Gender | Female (n=152,884) | 1.056 (0.969-1.150) | 1.079 (0.989-1.177) | 1.102 (1.004-1.210) | 1.126 (1.014-1.251) |
|  | Male (n=433,645) | 1.057 (1.006-1.111) | 1.091 (1.038-1.147) | 1.135 (1.075-1.198) | 1.186 (1.115-1.260) |
| Industry | Public Administration (n=179,388) | 1.067 (0.989-1.151) | 1.097 (1.015-1.185) | 1.129 (1.038-1.228) | 1.163 (1.057-1.279) |
|  | Transportation And Warehousing (n= 93,912) | 1.031 (0.916-1.161) | 1.066 (0.946-1.203) | 1.119 (0.984-1.272) | 1.186 (1.028-1.368) |
|  | Manufacturing (n= 82,406) | 1.057 (0.957-1.168) | 1.078 (0.973-1.193) | 1.096 (0.980-1.225) | 1.112 (0.977-1.265) |
|  | Accommodation And Food Services (n= 78,278) | 1.053 (0.930-1.193) | 1.076 (0.948-1.220) | 1.100 (0.961-1.259) | 1.125 (0.967-1.309) |
|  | Construction (n= 63,823) | 1.085 (0.945-1.246) | 1.128 (0.980-1.298) | 1.181 (1.015-1.374) | 1.242 (1.049-1.472) |
|  | Administrative And Support And Waste Management And Remediation (n= 47,843) | 1.064 (0.914-1.238) | 1.097 (0.940-1.279) | 1.135 (0.962-1.340) | 1.179 (0.978-1.421) |
|  | Wholesale Trade (n= 33,598) | 1.033 (0.868-1.229) | 1.068 (0.894-1.274) | 1.120 (0.924-1.357) | 1.186 (0.954-1.475) |
|  | Utilities (n= 5,305) | 1.025 (0.668-1.574) | 1.076 (0.695-1.667) | 1.165 (0.724-1.876) | 1.287 (0.747-2.219) |
|  | Agriculture, Forestry, Fishing And Hunting (n= 5,248) | 1.099 (0.752-1.605) | 1.171 (0.794-1.728) | 1.281 (0.833-1.969) | 1.424 (0.860-2.356) |
| Cause of Injury | Overexertion & bodily reaction (n=155,499) | 1.056 (0.975-1.143) | 1.080 (0.996-1.171) | 1.107 (1.014-1.208) | 1.134 (1.026-1.253) |
|  | Struck by/against (n=128,576) | 1.044 (0.953-1.144) | 1.080 (0.984-1.186) | 1.133 (1.025-1.253) | 1.199 (1.071-1.344) |
|  | Falls & slips (n=116,350) | 1.055 (0.959-1.161) | 1.070 (0.970-1.179) | 1.078 (0.970-1.197) | 1.081 (0.959-1.217) |
|  | Motor vehicle (n= 48,349) | 1.050 (0.887-1.242) | 1.085 (0.915-1.287) | 1.127 (0.939-1.352) | 1.174 (0.959-1.437) |
|  | Contact with objects/equipment (n= 32,409) | 1.008 (0.843-1.206) | 1.014 (0.845-1.217) | 1.027 (0.843-1.251) | 1.045 (0.835-1.307) |
|  | Other/miscellaneous (n= 29,916) | 1.098 (0.901-1.338) | 1.181 (0.966-1.444) | 1.309 (1.055-1.623) | 1.477 (1.161-1.880) |
|  | Burns & exposure (n= 27,166) | 1.112 (0.916-1.349) | 1.180 (0.969-1.437) | 1.270 (1.027-1.571) | 1.379 (1.085-1.754) |
|  | Cut/puncture/scrape (n= 26,530) | 1.144 (0.930-1.407) | 1.189 (0.963-1.468) | 1.227 (0.978-1.540) | 1.261 (0.977-1.627) |
|  | Caught in/between (n= 15,877) | 1.029 (0.804-1.317) | 1.043 (0.812-1.341) | 1.060 (0.807-1.391) | 1.078 (0.791-1.469) |
|  | Assault & violence (n= 10,585) | 1.083 (0.755-1.553) | 1.118 (0.776-1.612) | 1.159 (0.784-1.714) | 1.205 (0.780-1.861) |
| Nature of Injury | Traumatic musculoskeletal (n=398,811) | 1.045 (0.993-1.100) | 1.069 (1.015-1.126) | 1.098 (1.038-1.161) | 1.130 (1.060-1.204) |
|  | Open wounds & external trauma (n=122,229) | 1.082 (0.985-1.189) | 1.116 (1.014-1.227) | 1.153 (1.040-1.278) | 1.193 (1.062-1.340) |
|  | Multiple / unspecified (n= 61,697) | 1.062 (0.923-1.222) | 1.096 (0.950-1.263) | 1.137 (0.976-1.324) | 1.184 (0.998-1.405) |
|  | Concussion / head injury (n= 4,972) | 1.083 (0.697-1.683) | 1.092 (0.697-1.711) | 1.079 (0.663-1.756) | 1.053 (0.605-1.832) |
|  | Systemic / internal conditions (n= 3,548) | 1.872 (1.018-3.443) | 2.919 (1.576-5.405) | 5.212 (2.706-10.036) | 10.167 (4.928-20.977) |
| Length of Employment | <3 months (n= 47,575) | 1.064 (0.915-1.237) | 1.103 (0.946-1.285) | 1.154 (0.979-1.361) | 1.217 (1.011-1.464) |
|  | 3-12 months (n= 66,393) | 1.046 (0.931-1.176) | 1.079 (0.958-1.216) | 1.124 (0.989-1.279) | 1.179 (1.020-1.363) |
|  | 1-2 years (n= 46,639) | 1.033 (0.899-1.188) | 1.073 (0.932-1.237) | 1.138 (0.977-1.326) | 1.223 (1.029-1.455) |
|  | ≥2 years (n=221,791) | 1.053 (0.983-1.129) | 1.073 (1.000-1.152) | 1.092 (1.013-1.178) | 1.111 (1.020-1.209) |
|  | Missing (n=208,859) | 1.066 (0.993-1.144) | 1.102 (1.025-1.184) | 1.145 (1.060-1.237) | 1.194 (1.095-1.302) |
| Time of Injury | Morning (6am-12pm) (n=157,005) | 1.056 (0.973-1.146) | 1.079 (0.993-1.173) | 1.104 (1.009-1.207) | 1.129 (1.021-1.248) |
|  | Afternoon (12pm-6pm) (n=142,470) | 1.057 (0.969-1.154) | 1.091 (0.998-1.192) | 1.132 (1.029-1.246) | 1.180 (1.061-1.314) |
|  | Evening/Night (6pm-6am) (n=117,592) | 1.043 (0.954-1.141) | 1.069 (0.976-1.171) | 1.103 (1.000-1.216) | 1.142 (1.023-1.276) |
|  | Missing (n=174,190) | 1.065 (0.988-1.148) | 1.101 (1.020-1.189) | 1.147 (1.056-1.246) | 1.200 (1.093-1.317) |
| Region | Downstate (n=335,701) | 1.048 (1.007-1.091) | 1.079 (1.036-1.124) | 1.120 (1.071-1.171) | 1.169 (1.110-1.230) |
|  | Upstate (n=255,556) | 1.058 (1.003-1.115) | 1.080 (1.024-1.140) | 1.103 (1.041-1.169) | 1.126 (1.054-1.203) |

**Source:** Authors' analysis of New York State Workers' Compensation Board claims data, 2016-2024, and daily meteorological data from GridMET.

**Notes:** Odds ratios were estimated from time-stratified case-crossover analyses using conditional logistic regression with natural cubic splines (3 degrees of freedom), adjusted for precipitation. Effect modification was assessed using stratified models.

### Supplemental Figure 1. Sensitivity by analytic sample

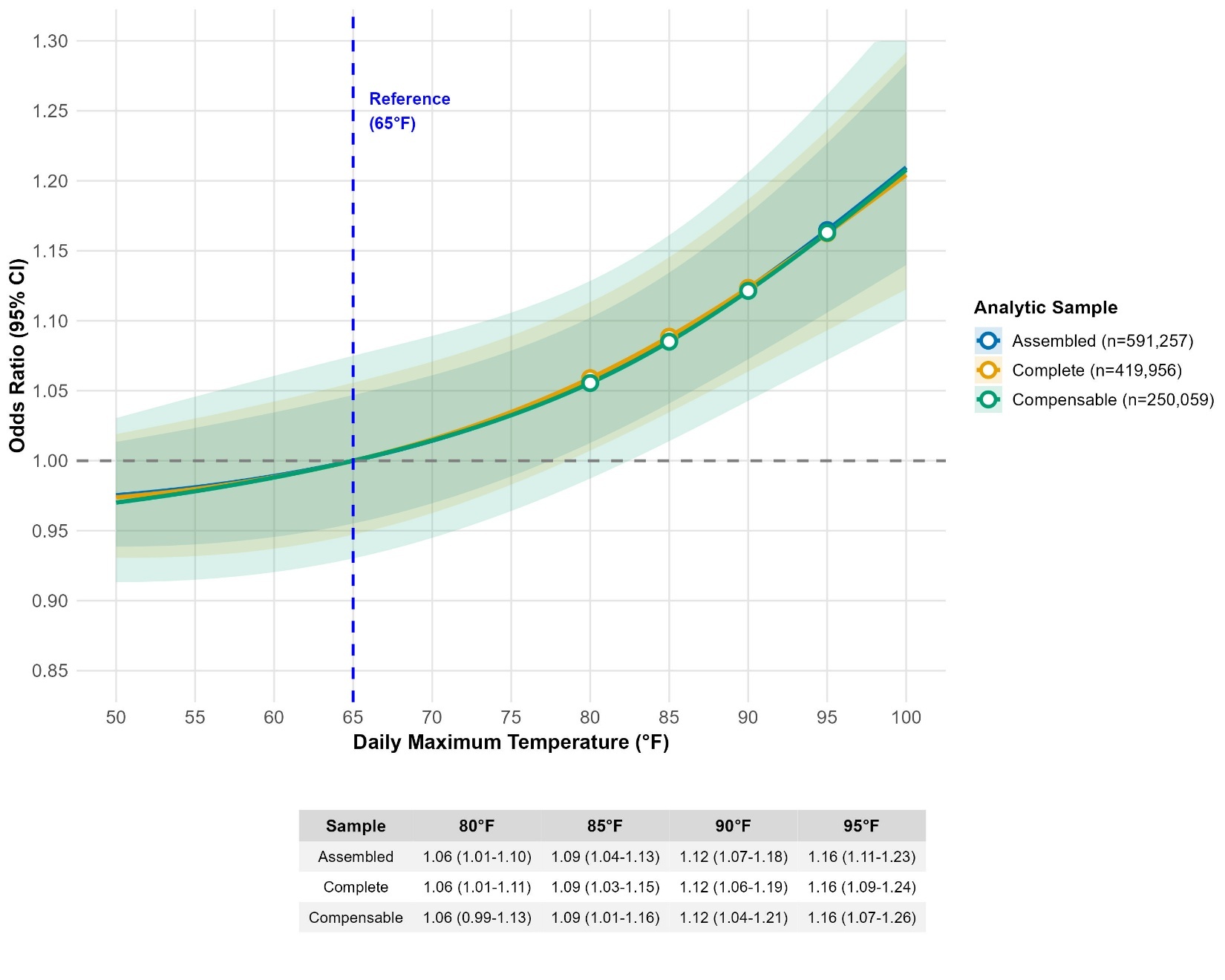

**Source:** Authors' analysis of New York State Workers' Compensation Board claims data, 2016-2024, and daily meteorological data from GridMET. **Notes:** Odds ratios and 95% confidence intervals (shaded areas) were estimated from time-stratified case-crossover analyses using conditional logistic regression with natural cubic splines (3 degrees of freedom), adjusted for precipitation. Assembled claims include all work-related injuries and illnesses for which the Board opened a case file, regardless of compensability. Complete claims are assembled claims with formal notice of injury from the insurer and qualifying medical documentation, while established claims are a subset of complete claims for which the Board determined work-relatedness and employer liability.

### Supplemental Figure 2. Sensitivity using heat index

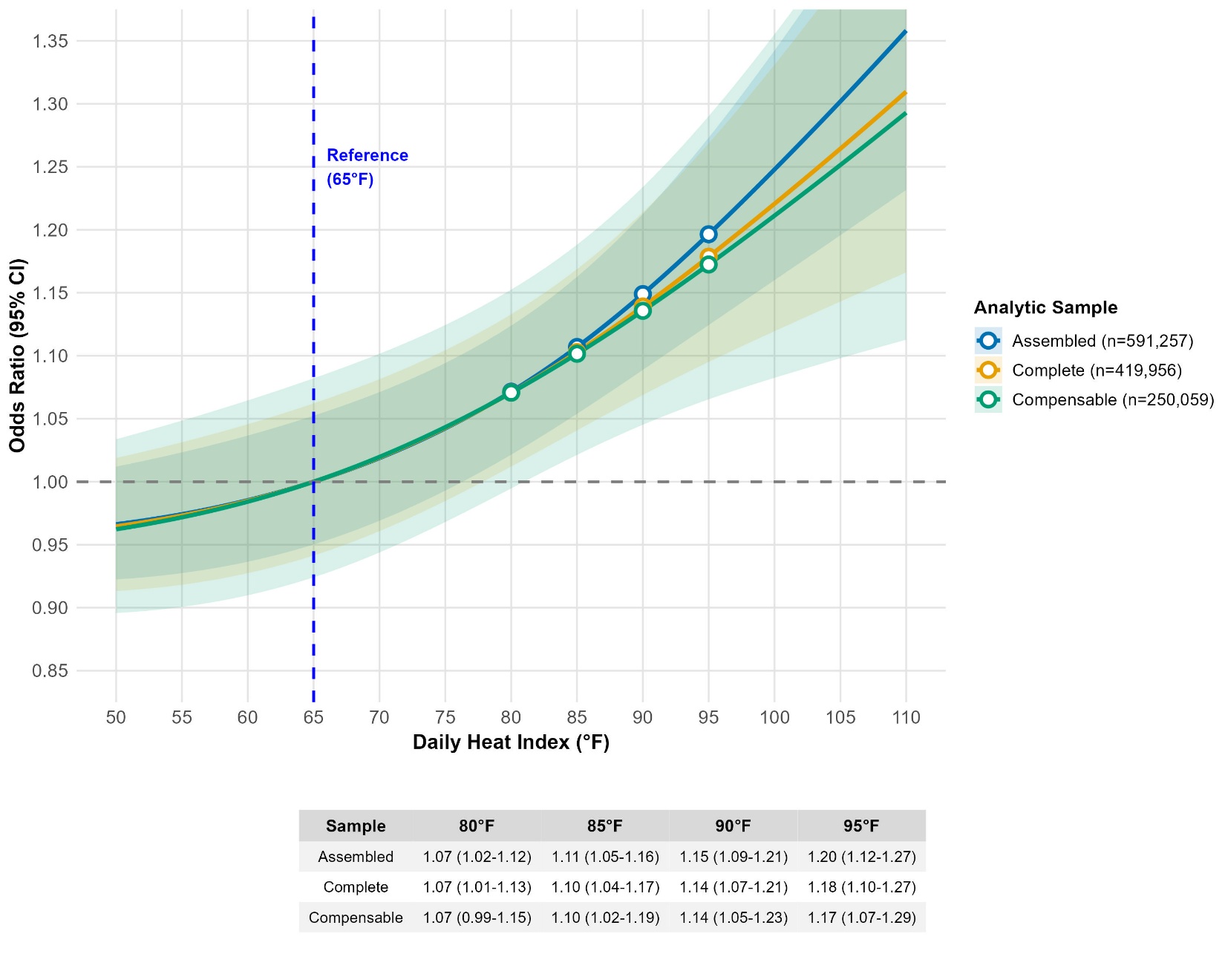

**Source:** Authors' analysis of New York State Workers' Compensation Board claims data, 2016-2024, and daily meteorological data from GridMET. **Notes:** Odds ratios and 95% confidence intervals (shaded areas) were estimated from time-stratified case-crossover analyses using conditional logistic regression with natural cubic splines (3 degrees of freedom), adjusted for precipitation. Heat index was calculated using daily maximum temperature and mean daily relative humidity and evaluated as a sensitivity analysis. Assembled claims include all work-related injuries and illnesses for which the Board opened a case file, regardless of compensability. Complete claims are assembled claims with formal notice of injury from the insurer and qualifying medical documentation, while established claims are a subset of complete claims for which the Board determined work-relatedness and employer liability.

### Supplemental Figure 3: Odds ratios for temperature-injury relationship by age group

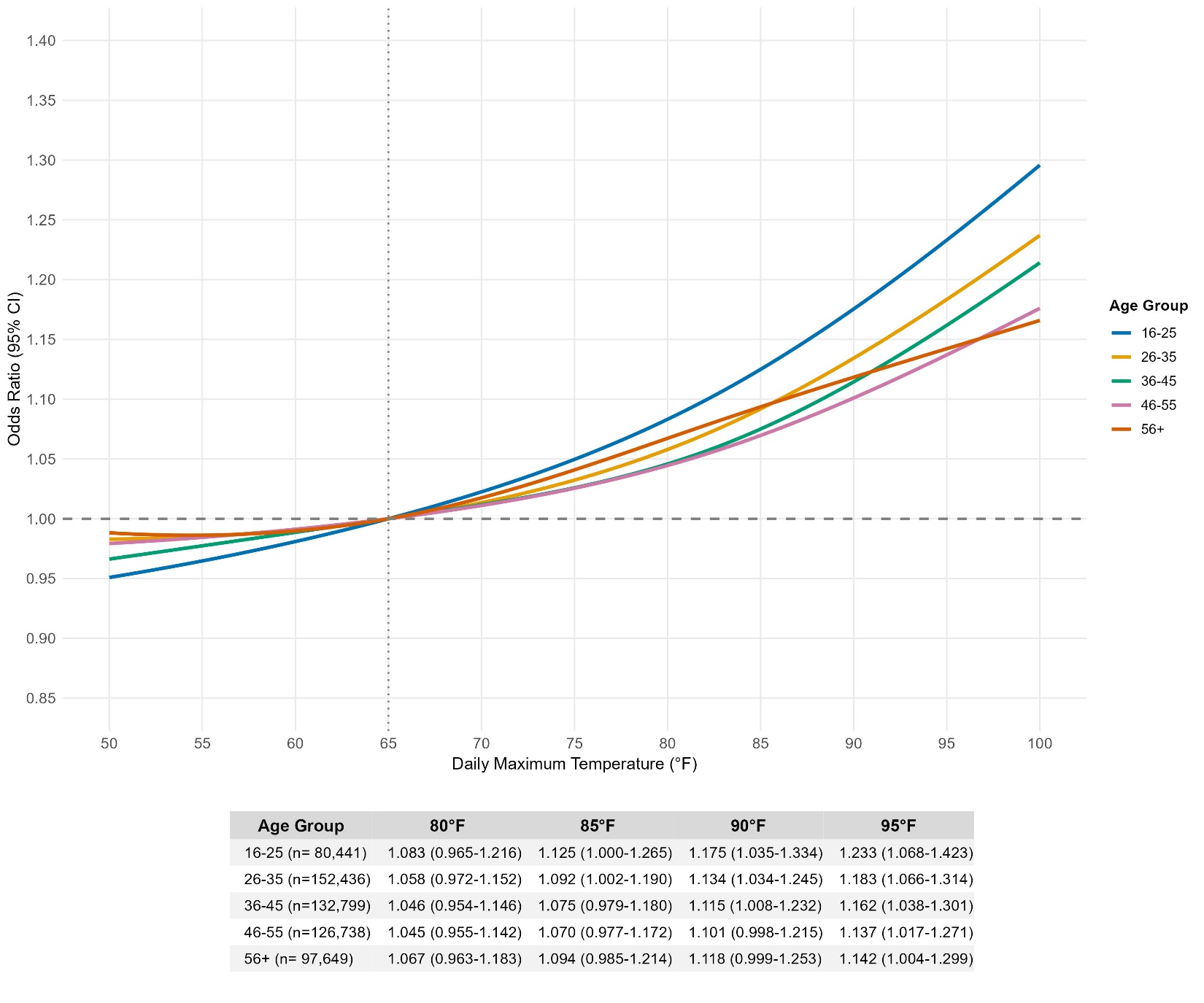

**Source:** Authors' analysis of New York State Workers' Compensation Board claims data, 2016-2024, and daily meteorological data from GridMET. **Notes:** Odds ratios and 95% confidence intervals (shaded areas) from time-stratified case-crossover analyses using conditional logistic regression with natural cubic splines (3 degrees of freedom), adjusted for precipitation. Reference temperature is 65°F.

### Supplemental Figure 4: Odds ratios for temperature-injury relationship by gender

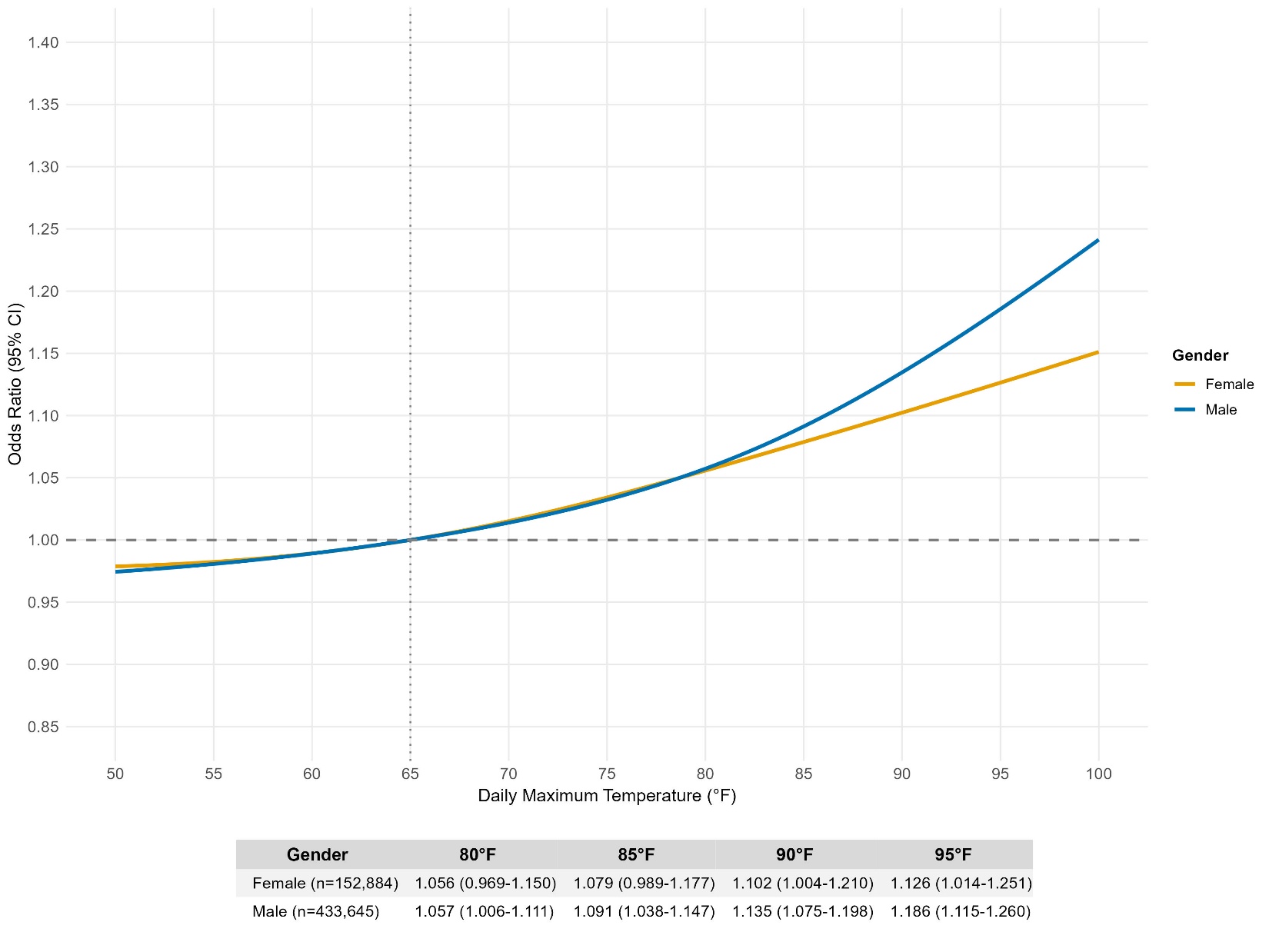

**Source:** Authors' analysis of New York State Workers' Compensation Board claims data, 2016-2024, and daily meteorological data from GridMET. **Notes:** Odds ratios and 95% confidence intervals (shaded areas) from time-stratified case-crossover analyses using conditional logistic regression with natural cubic splines (3 degrees of freedom), adjusted for precipitation. Reference temperature is 65°F.

### Supplemental Figure 5: Odds ratios for temperature-injury dose-relationship by length of employment

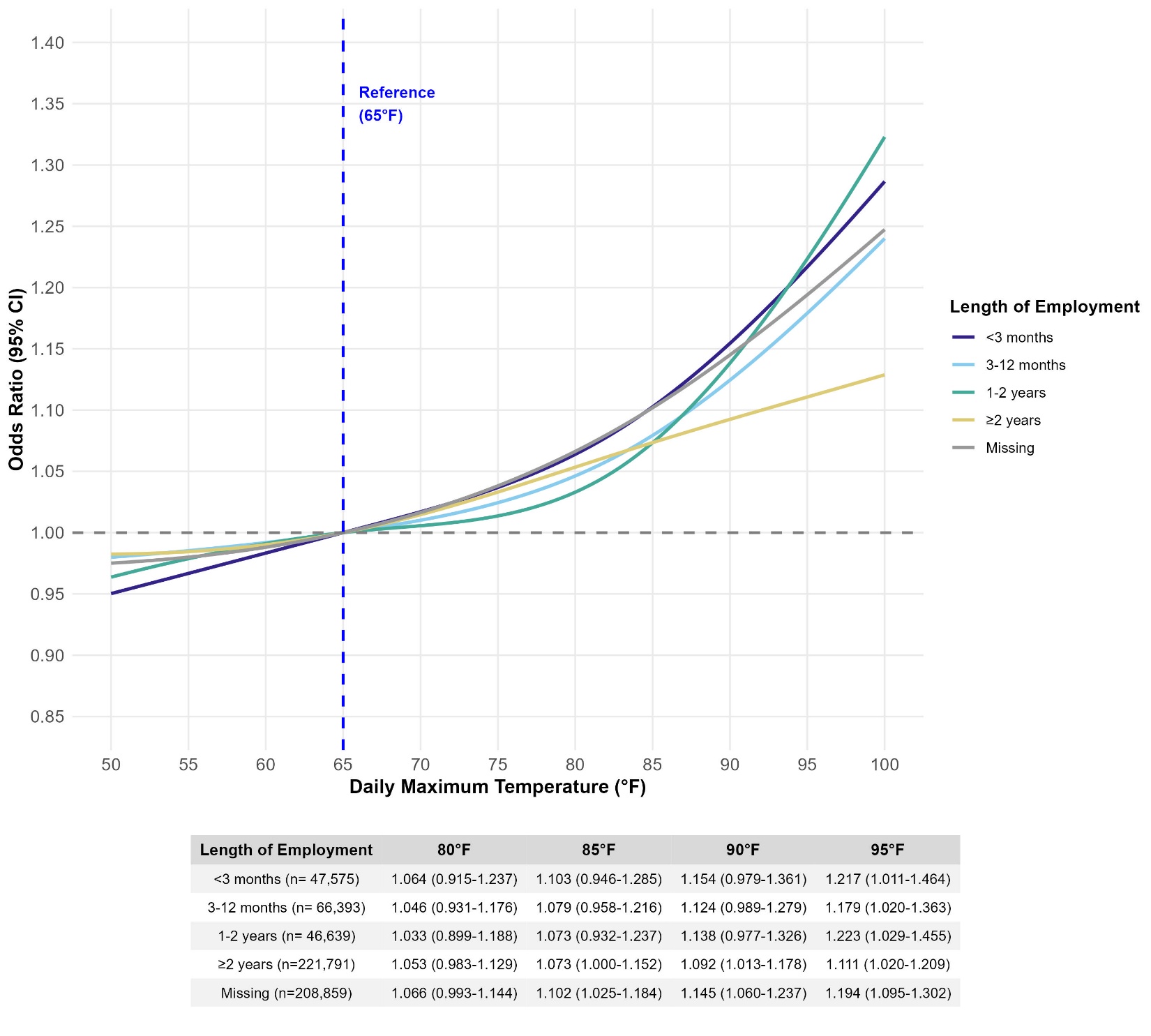

**Source:** Authors' analysis of New York State Workers' Compensation Board claims data, 2016-2024, and daily meteorological data from GridMET. **Notes:** Odds ratios and 95% confidence intervals (shaded areas) from time-stratified case-crossover analyses using conditional logistic regression with natural cubic splines (3 degrees of freedom), adjusted for precipitation. Reference temperature is 65°F.

### Supplemental Figure 6: Odds ratios for temperature-Injury dose-response relationship by time of injury

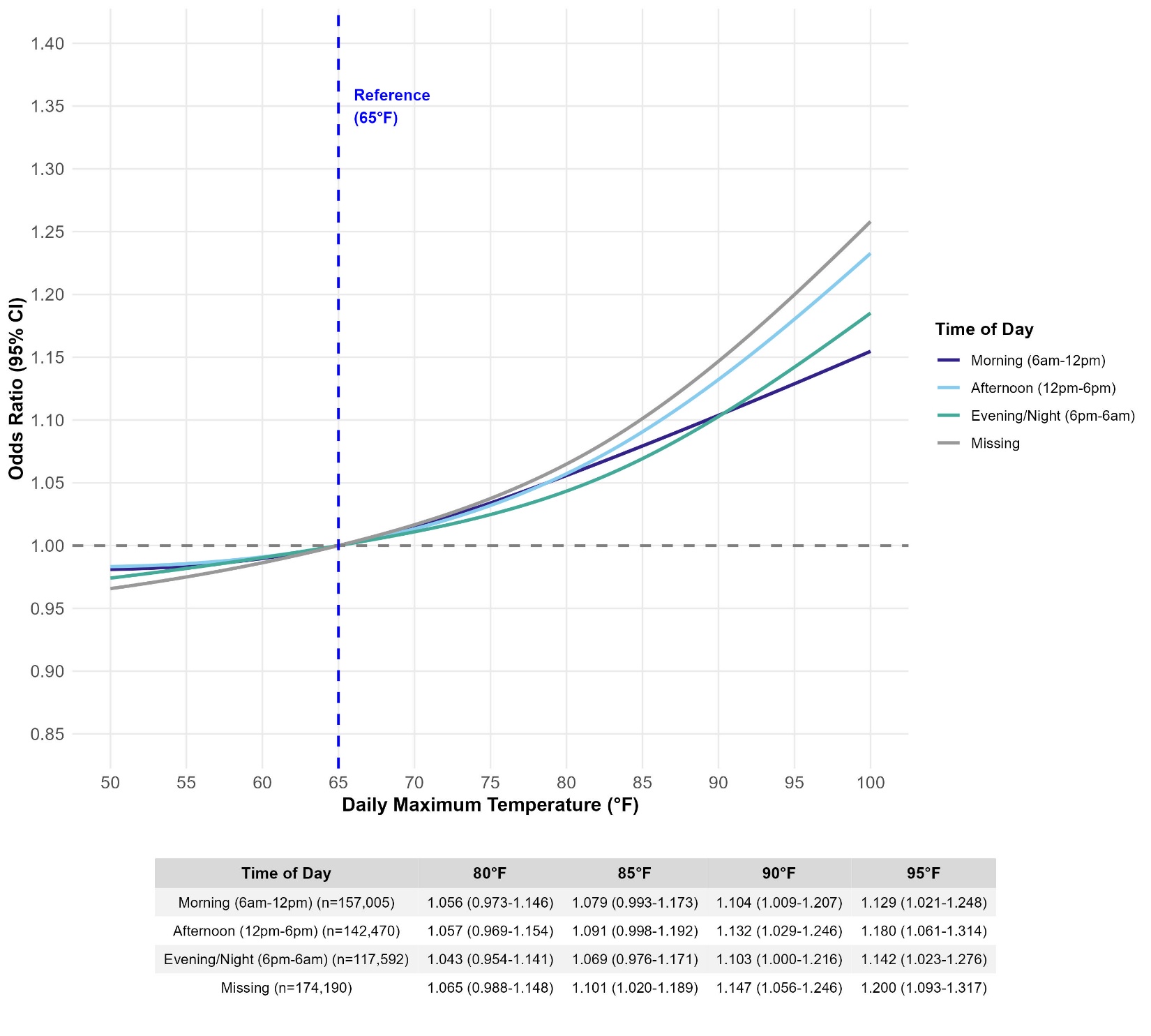

**Source:** Authors' analysis of New York State Workers' Compensation Board claims data, 2016-2024, and daily meteorological data from GridMET. **Notes:** Odds ratios and 95% confidence intervals (shaded areas) from time-stratified case-crossover analyses using conditional logistic regression with natural cubic splines (3 degrees of freedom), adjusted for precipitation. Reference temperature is 65°F.

### Supplemental Figure 7: Odds ratios for temperature-Injury dose-response relationship by region

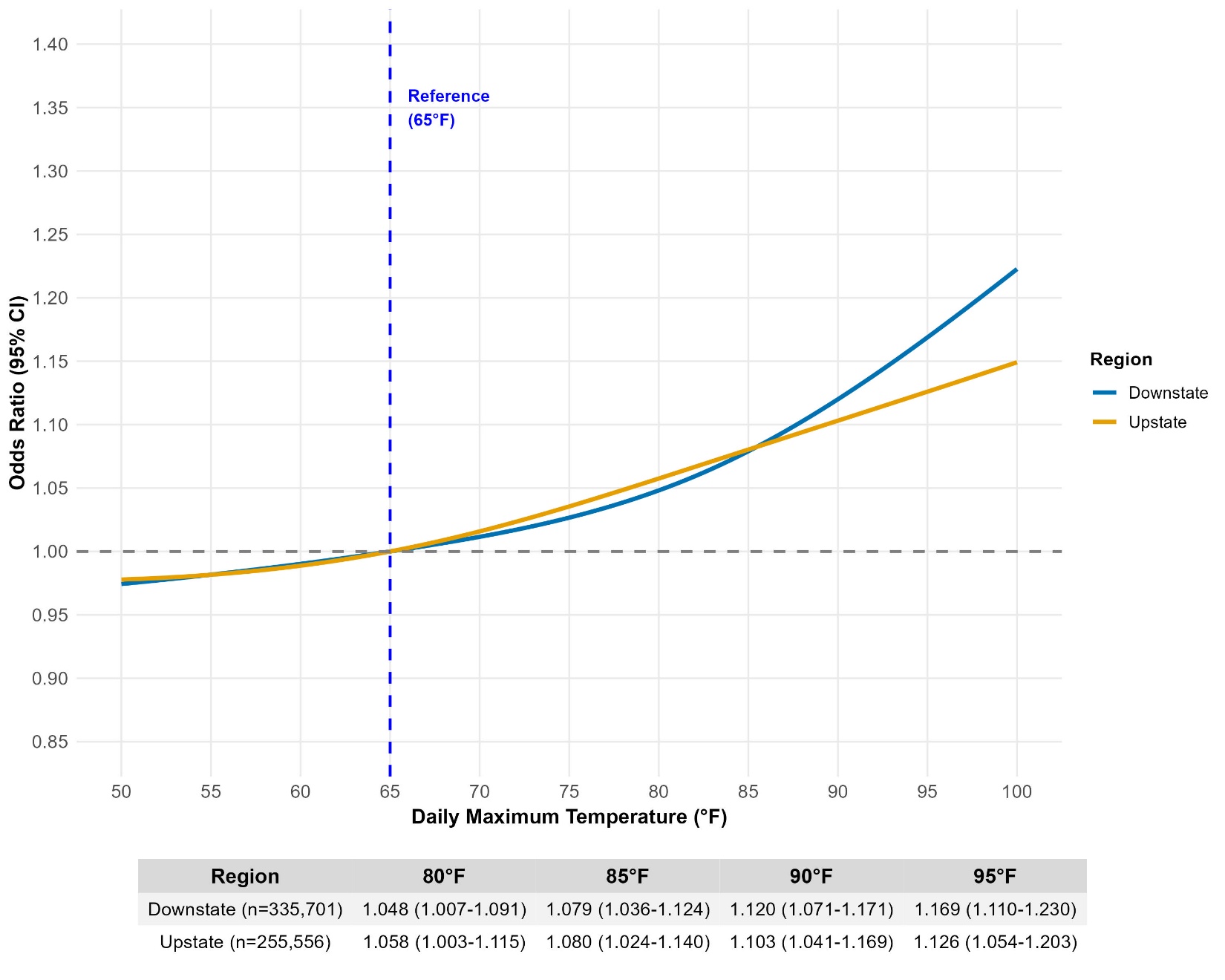

**Source:** Authors' analysis of New York State Workers' Compensation Board claims data, 2016-2024, and daily meteorological data from GridMET. **Notes:** Odds ratios and 95% confidence intervals (shaded areas) from time-stratified case-crossover analyses using conditional logistic regression with natural cubic splines (3 degrees of freedom), adjusted for precipitation. Reference temperature is 65°F.
